# Non-invasive spinal cord neuromodulation enables volitional anti-gravity leg movements after motor-complete spinal cord injury: responders vs. non-responders

**DOI:** 10.1101/2025.09.11.25335523

**Authors:** Raza N. Malik, Soshi Samejima, Alison MM Williams, Ali Hosseinzadeh, Emmanuel Ogalo, Alex Stolz, Chantal Lam, Claire Shackleton, Tiev Miller, Mohamed Gomaa Sobeeh, John LK Kramer, Tania Lam, Rahul Sachdeva, Michael J. Berger, Andrei Krassioukov

**Author notes:** **Correspondence.** Raza N. Malik,; Andrei V. Krassioukov.

## Abstract

**Background and Objectives:** Transcutaneous spinal cord stimulation (tSCS) is an emerging treatment for motor recovery following spinal cord injury (SCI). However, the extent of motor recovery with tSCS and the reasons why some individuals with motor-complete SCI respond less effectively, despite having the same injury classification, remain unclear. Here, we demonstrate that lumbosacral tSCS can enable anti-gravity voluntary movement following motor-complete SCI, and identify markers that distinguish responders from non-responders.

**Methods:** Ten individuals with chronic motor-complete SCI received 30Hz lumbosacral tSCS for 60 min, 2-5 times per week, for a minimum of 6 weeks. Post-intervention, volitional movement was measured using surface electromyography (EMG) over the quadriceps and tibialis anterior (TA), and knee and ankle joint range of motion. To identify markers of responsiveness, we assessed the integrity of the corticospinal tract (motor evoked potentials; MEPs), ascending sensory pathways (somatosensory evoked potentials; SEPs), spinal cord reflexes (H-reflex), and motor neurons (compound muscle action potential, CMAP), along with muscle morphology using ultrasound echo-intensity.

**Results:** Five of 10 individuals demonstrated voluntary anti-gravity knee extension and ankle dorsiflexion strength in the presence of tSCS. TA MEPs were observed in one responder only and tibial nerve SEPs were not observed in any participants. All participants showed poor TA muscle morphology. Four responders had a soleus H-reflex (compared to 2/5 non-responders) and a normal amplitude fibular CMAPs (compared to 2/5 non-responders).

**Discussion:** These results show that tSCS can enable volitional motor activity against gravity in people with motor-complete SCI, but there is variability in responsiveness. Using conventional neurophysiological techniques, we were unable to consistently demonstrate the pathways facilitating voluntary control or the factors differentiating responders versus non-responders, but trends were observed. Spinal cord reflex and peripheral motor nerve integrity may be important for responding to tSCS but may not distinguish responders from non-responders. Additional assessments are needed to develop biomarkers for stratifying motor responders to tSCS.

## Introduction

Spinal cord injury (SCI) damages crucial spinal circuits leading to impaired voluntary motor control. Many individuals with clinically complete SCI have spared neural connections across the lesion site, but that are unable to generate functional movement.^1^ Transcutaneous spinal cord stimulation (tSCS) may engage these spared circuits to enable voluntary lower limb movements after SCI.^2–4^ Previous studies primarily show gravity-eliminated volitional lower extremity movements with tSCS in those with chronic motor-complete SCI,^2–4^ but gravity-eliminated movements may not translate to improved function and recovery of volitional motor control against gravity remains elusive.^3^

The inter-individual variability in responsiveness to electrical neuromodulation has been demonstrated^5,6^ and the factors influencing this variability have yet to be investigated. Through identification of markers of responsiveness to SCS, we can identify those that are most likely to regain motor function. Distinguishing characteristics in responders and non-responders to tSCS could also allow for understanding the underlying mechanisms of tSCS and improve the precision of treatment.^7^

Neurophysiological and imaging techniques, such as brain, peripheral nerve stimulation, and muscle ultrasound, are commonly used to assess descending motor pathways, spinal reflexes, lower motoneuron integrity, and muscle quality – all important components in the generation of volitional movements. In this case series, we assessed voluntary anti-gravity leg movements in individuals with motor-complete SCI following tSCS therapy and also explored potential neurophysiological and anatomical markers of tSCS responsiveness. We hypothesized that corticospinal integrity and peripheral motor nerve integrity could differentiate between motor responders and non-responders to tSCS therapy.

## Methods

Ten individuals (44.3 ± 9.8 years of age) with traumatic, chronic (average time since injury of 13.15 ± 11.5 years), motor-complete SCI (American Spinal Cord Injury Association Impairment Scale (AIS A or B)), above T6, were identified from three ongoing clinical trials registered on ClinicalTrials.gov: NCT04726059 (first enrolled: March 22, 2021), NCT04604951 (first enrolled: May 4, 2021), and NCT05369520 (first enrolled: October 3, 2022).^8,9^ Participants met the specific inclusion criteria for their respective trials (e.g., Chronic motor-complete SCI above T6). During the course of these trials, we observed the return of seated voluntary lower limb motor activity in the presence of tSCS in half of the individuals (responders). In these individuals the common movements regained were manual muscle strength grade 3 or higher (i.e., antigravity) knee extension and ankle dorsiflexion. Video analysis of voluntary movements for each participant are presented in Videos 1-5 (preprint: videos available upon request from the corresponding author). The first signs of volitional control occurred at different times for each responder, with a median of 7 sessions (range: 4-12). In the other half of the participants, we did not observe the return of voluntary movements (non-responders). Non-responders were chosen to best match the responders age and injury level from the same clinical trials.

We used the International Standards for Neurological Classification of Spinal Cord Injury (ISNCSCI) to determine neurological level of injury (NLI) and AIS grade.^10^ A summary of participant demographics and injury characteristics is provided in Table 1. The trials and analyses used in this manuscript were approved by the University of British Columbia’s Clinical Research Ethics Board (H20-01307, H22-00365, H20-01163, H22-03727), and was conducted in accordance with the Declaration of Helsinki. All participants provided written informed consent prior to participation and completed a signed consent-to-disclose form for the use of their data and videos in this study.

**Table 1.** Demographics of responders and non-responders.

| ID | Age-range (yrs) | Sex | AIS | NLI | Lumbosacral tSCS @ 30Hz with 10kHz carrier |
| --- | --- | --- | --- | --- | --- |
| R01 | 20-30 | M | B | C4 | 5x/wk for 6wks |
| R02 | 40-50 | M | A | T5 | 2x/wk for 6wks |
| R03 | 40-50 | M | A | T6 | 3x/wk + BWSTT for 12 wks |
| R04 | 40-50 | M | A | T5 | 3x/wk + BWSTT for 12 wks |
| R05 | 50-60 | M | B | C5 | 3x/wk for 8wks |
| NR01 | 30-40 | F | A | C4 | 2x/wk for 6wks |
| NR02 | 50-60 | M | A | T5 | 5x/wk for 6 wks |
| NR03 | 50-60 | M | A | C7 | 5x/wk for 6 wks |
| NR04 | 50-60 | F | A | T5 | 2x/wk for 6wks |
| NR05 | 30-40 | M | A | T2 | 3x/wk for 8 wks |

**Abbreviations:** R, Responder; NR, Non-Responder; AIS, American Spinal Injury Association Impairment Scale; NLI, Neurological Level of Injury; TSI, Time Since Injury; tSCS, Transcutaneous Spinal Cord Stimulation, NA, Not applicable. BWSTT, body weight supported treadmill training. Note: The International Standards for Neurological Classification of Spinal Cord Injury (ISNCSCI) was used to determine neurological level of injury (NLI) and AIS grade

### Intervention and Protocol

Reporting of the SCS protocol and intervention adhered to minimum reporting standards.^11^ Participants received lumbosacral tSCS 2-5 times per week for 6-12 weeks depending on the clinical trial they were enrolled in. Charge-balanced monophasic 1ms pulses at 30Hz with a 10kHz carrier frequency were used. tSCS intensity was set based on comfort, with all participants tolerating the maximum 130mA. Briefly, those enrolled in the “Below the Belt” trial [NCT04726059] (R01, R02, NR01, NR02, NR03, NR04, see Table 1), received 60 min sessions 2x or 5x/week stimulation in a seated position, in the absence of any activity-based therapy.^8^ Those enrolled in the “MACHINE” Trial [NCT04604951] (R03 and R04) received TSCS 3x/week concurrently with 45 min of robotic locomotor training on the bodyweight-supported treadmill using the Lokomat system.^9^ Lastly, two participants (R05 and NR05) enrolled in NCT05369520 received 60 min sessions 3x/week stimulation targeting the lumbosacral spinal cord in a seated position, in the absence of any activity-based therapy.

tSCS was administered using a non-invasive central nervous system stimulator (TESCoN, SpineX Inc., Northridge, CA). Stimulation was delivered via a 3.2 cm standard round self-adhesive electrode (ValuTrode, Axelgaard Manufacturing Co., Ltd, Fallbrook, CA) placed on the midline below the T11 and L1 spinous processes as the cathode electrodes. Two 5 ×10 cm rectangular electrodes (ValuTrode, Axelgaard Manufacturing Co., Ltd, Fallbrook, CA), placed symmetrically over the skin of the iliac crests, served as the anode electrodes. This electrode montage targeted the lumbosacral spinal cord segments.

### Post-intervention behavioural, neurophysiological, and imaging assessments and analysis

To quantify voluntary muscle activity and movement in the lower limbs post-intervention, we recorded surface electromyography (EMG) and kinematic data as participants performed three cycles of 4-second contractions followed by 4 seconds of relaxation, involving simultaneous voluntary knee extension and ankle dorsiflexion with and without real-time tSCS (see *Assessments and Analysis* below). Responders were defined as individuals capable of performing knee extension and ankle dorsiflexion from a seated position, confirmed visually (See Videos 1-5 showing voluntary movements for each participant), whereas non-responders were unable to perform this movement.

We then conducted several neurophysiological and imaging assessments to identify biomarkers of responsiveness. For all assessments, we recorded data from the limb that demonstrated the highest strength grade in responders and in the non-responders we collected data from their dominant leg prior to injury.

#### Lower limb kinematics and EMG

We calculated the average change in hip, knee, and ankle angle from rest to maximum during knee extension and dorsiflexion across three contraction attempts. For each trial (with and without tSCS) and muscle, a threshold from background EMG at rest + 3 standard deviations (SD) was generated and change in muscle activity from rest to contraction was calculated by expressing the number of SDs above the threshold. Sagittal-plane kinematics of the hip, knee, and ankle joints were recorded at 100Hz using the Optotrak motion capture system (Northern Digital Inc, Waterloo, ON, Canada), by placing infrared emitting diodes over the greater trochanter, lateral epicondyle of the knee, lateral malleolus, and 5th metatarsal. EMG of the rectus femoris (RF), vastus lateralis (VL), biceps femoris (BF), tibialis anterior (TA) and soleus (SOL) were recorded at 2000Hz (Delsys Inc., Natick, USA). Kinematics and EMG were analyzed offline using custom MATLAB routines (Mathworks Inc., Natick, MA, USA).

Kinematics were filtered using a low-pass 4th order Butterworth filter with a cut-off frequency of 6Hz. EMG data were offset and were filtered using a bandpass 4th order Butterworth filter with cut-off frequencies of 10-100Hz. Notch Filters at harmonics of 30Hz were also used to reduce the noise in the signal introduced by transcutaneous spinal cord stimulation (tSCS). Finally, a root mean square (RMS) envelope was applied to the EMG data with an RMS window size of 200.

##### Motor evoked potentials

We evaluated the integrity of descending motor pathways by assessing the presence of motor-evoked potentials (MEPs) in the TA, induced by transcranial magnetic stimulation (TMS) over the leg area of the primary motor cortex.^12^ For this tests, each participant rested in an upright, seated position in their wheelchair or, at their request for comfort, in the supine position on a plinth. Stimulation was delivered using a 110 cm double cone coil (Magstim Rapid Pulse2, Magstim Company, UK). EMG recordings were taken from the TA (Delsys Inc., Natick, USA). EMG recordings were made at 2000Hz and were plotted in real-time for visual inspection, and stored for offline analysis. Surface anatomy was used to locate the anticipated optimal point of excitability of the primary motor cortex location of the TA.^13^ We attempted to verify the location and elicit an MEP for each participant by stimulating at approximately 80% of the maximum stimulator output (%MSO) in a 5×5cm grid around the anticipated hot spot location. We delivered blocks of stimuli at increasing intensity until a consistent MEP response was elicited, or, if a MEP was not elicited, until the maximum stimulator output. As participants were unable to contract their TA to provide facilitating background EMG, we asked them to perform the Jendrassik maneuver in all trials. This involved interlocking their fingers, clenching their jaw, and attempting to pull their fingers apart (participants with tetraplegia who were unable to interlock their fingers were asked to only clench their jaw and tense their upper body). If an MEP was elicited, we attempted to generate a recruitment curve (input-output curve) of the responses in both conditions. We delivered blocks of 5-10 pulses beginning below motor threshold and increased in intensity by 5-10% MSO for each block. The test ended when 100% MSO was reached, the MEP amplitude reached a plateau, or the participant requested to discontinue increasing stimulation intensity.

We used custom-written MATLAB Scripts (Mathworks Inc., Natick, MA, USA) to analyze MEP responses. For each MEP response, a 100ms window of the rectified EMG signal 50ms before the TMS pulse was averaged to define a threshold for MEP presence. If the EMG data after stimulation exceeded two standard deviations of this background window, and surpassed the threshold for at least 2ms, then the MEP was considered present.^14,15^ All responses were verified visually by a member of the research team with experience in MEP analysis in SCI motor-complete participants. The MEP latency was defined as the instance at which the EMG first crossed the threshold. We then measured the peak-to-peak amplitude of the TA muscle EMG signal after MEP latency. Peak-to-peak MEP amplitudes at the same %MSO were then averaged and plotted against stimulation intensity to produce a recruitment curve.

##### Somatosensory evoked potentials

We evaluated the integrity of ascending sensory pathways by evaluating somatosensory evoked potentials (SEPs).^12^ For this procedure, stimulating surface electrodes were placed over the skin on the posterior tibial nerve behind the medial malleolus. Participants also wore a cap fitted with a 32-channel electroencephalography (EEG) system in the international 10-20 system (Brain Products GmbH, Gilching, Germany). Gel was injected into the space below each electrode to ensure adequate impedance and electrode signal quality. Tibial nerve SEPs were elicited using monophasic square-wave electrical pulses (1 ms duration, delivered at 1-3 Hz). For the intensity of stimulation, we first attempted to find the perceptual threshold, and if we did, we stimulated at 3 times the perceptual threshold. If the perceptual threshold could not be found due to SCI, then we found motor threshold (intensity at which a motor response was observed in the foot) and stimulated at 1-2 times the motor threshold. For testing, participants were seated in their wheelchair with eyes closed and instructed to remain relaxed in a quiet environment. EEG data were recorded at 1000 Hz and data from the Cz electrode were analyzed, referenced to the Fz electrode. We processed EEG data using custom MATLAB scripts (Mathworks Inc., Natick, MA, USA) utilizing functions from the EEGlab toolbox (V.14_1_1b, SCCN). We filtered EEG data using a bandpass 2nd order Butterworth filter with a cut off frequency of 1-100 Hz. We then divided the EEG data into epochs, each sectioned from 500 ms pre-stimulus to 1000 ms post-stimulus and baseline corrected. A total of 600 evoked potential trials were averaged and visually inspected for the presence of a P40-N50 SEP waveform. If present, the peak-to-peak amplitude and latencies were extracted.

##### H-reflex

We evaluated spinal cord circuit integrity using the soleus Hoffman reflex (H-reflex).^16,17^ For this assessment, participants were in an upright seated position with their foot partially elevated and knee extended. Stimulating surface electrodes were placed in the popliteal fossa over the tibial nerve with the anode distal to the cathode. The EMG electrodes (Delsys Inc., Natick, USA) were placed on the ipsilateral soleus (SOL) muscle, below the gastrocnemius, to record H-Reflex and M-Wave responses to tibial nerve stimulation. EMG recordings were made at 2000Hz, plotted in real-time for visual inspection, and stored for offline analysis using custom written MATLAB scripts (Mathworks Inc., Natick, MA, USA). To generate an H-reflex recruitment curve (input-output curve), 1ms square-wave pulses with an inter-pulse period of 5 seconds were delivered to the tibial nerve starting at sub-threshold intensities for eliciting an H-reflex (DS7 Constant Currant Stimulator, Digitimer, Ltd., UK). Stimulations of progressively higher intensity were delivered until a plateau in the peak-to-peak amplitude of the M-Wave was observed. For each trial, the peak-to-peak amplitude of the M-wave and H-Reflex were extracted. The H_max_/M_max_ ratio was then calculated to represent the percentage of the motoneuron pool that could be activated during stimulation.^18^

##### Compound muscle action potential

We evaluated peripheral motor nerve integrity by assessing compound muscle action potentials (CMAPs) from stimulating the fibular nerve at the fibular head and recording over TA by delivering single pulses (pulse width 0.2-0.5 ms, current 10-100 mA).^19^ The intensity of the current was incrementally increased until a plateau in the response potential was observed. Once a plateau was observed, the stimulus duration was increased to determine if any additional response was seen. Each response was visually inspected to ensure there was no initial positive dip or change in waveform morphology that would be indicative of co-stimulation. CMAPs were recorded and analyzed using a commercially available EMG machine (Dantec Keypoint G4). We extracted the negative peak amplitude of the CMAP. CMAPs were considered normal if they were ≥ 3.0 mV. ^20^

##### Ultrasound muscle morphology

We assessed muscle morphology of the TA using B-mode ultrasound (MyLab Alpha, ESAOTE S.P.A, Genova, Italy) with a 2-12 MHz linear array transducer^21,22,23^. Images were acquired by a board-certified physiatrist with 10 years of muscle imaging experience. For this procedure, a 1.5-2mm thick gel coupling layer served as the interface between the skin surface and transducer, which was placed on the TA muscle belly approximately one third the distance between the lateral malleolus and the fibula head. To ensure minimal anisotropy during contact, the transducer was angled caudally and cranially until the fascial borders were optimized as distinct from muscle tissue. Minimal pressure and copious gel were used to reduce deformation of the muscle. Images were obtained and exported for post-processing and analysis. Open source software (ImageJ version IJ 1.46r, National Institutes of Health, USA) was used to identify the region of interest (ROI) (i.e., fascial boundaries of the TA for each cross-sectional image), and generate an echo-intensity value for pixels within the ROI.^23^ The darkest and lightest pixels represent upper and lower value limits ranging from 0 to 255, respectively. Higher values generally indicate infiltration of intramuscular adiposity, fibrosis, greater organizational density of collagen, reduced contractile capacity and are associated with functional impairment and neuromuscular disease progression.^23,24^ To obtain normative estimates of echogenicity, we collected ultrasound data from 20 healthy controls (12 males and 8 females, mean age 39.5 years [range: 23-61]). The mean normative echo-intensity value of the TA was 50.5, with a 95% confidence interval of 45.8-55.1.

## Results

Following the multi-week intervention, five individuals demonstrated volitional lower limb control in the presence of tSCS (responders), while five individuals did not show any motor activity (non-responders). The responders were all male, two were classified as AIS B and three were AIS A, with an average age of 42. ± 8.4 years, while the non-responders consisted of three males and two females, all were AIS A, with an average age of 46.2±11.8 years (Range 31-56; Table 1)

In responders, movements regained included anti-gravity knee extension and ankle dorsiflexion after the long-term tSCS intervention. Figure 1A shows attempted contraction and relaxation of combined knee extension and ankle dorsiflexion in a representative responder and non-responder with and without tSCS in a seated position (*see Figure 1B and C for kinematic and EMG quantifications*). Videos 1-5 show each responders’ attempts (preprint: videos available upon request from the corresponding author).

**Figure 1.**
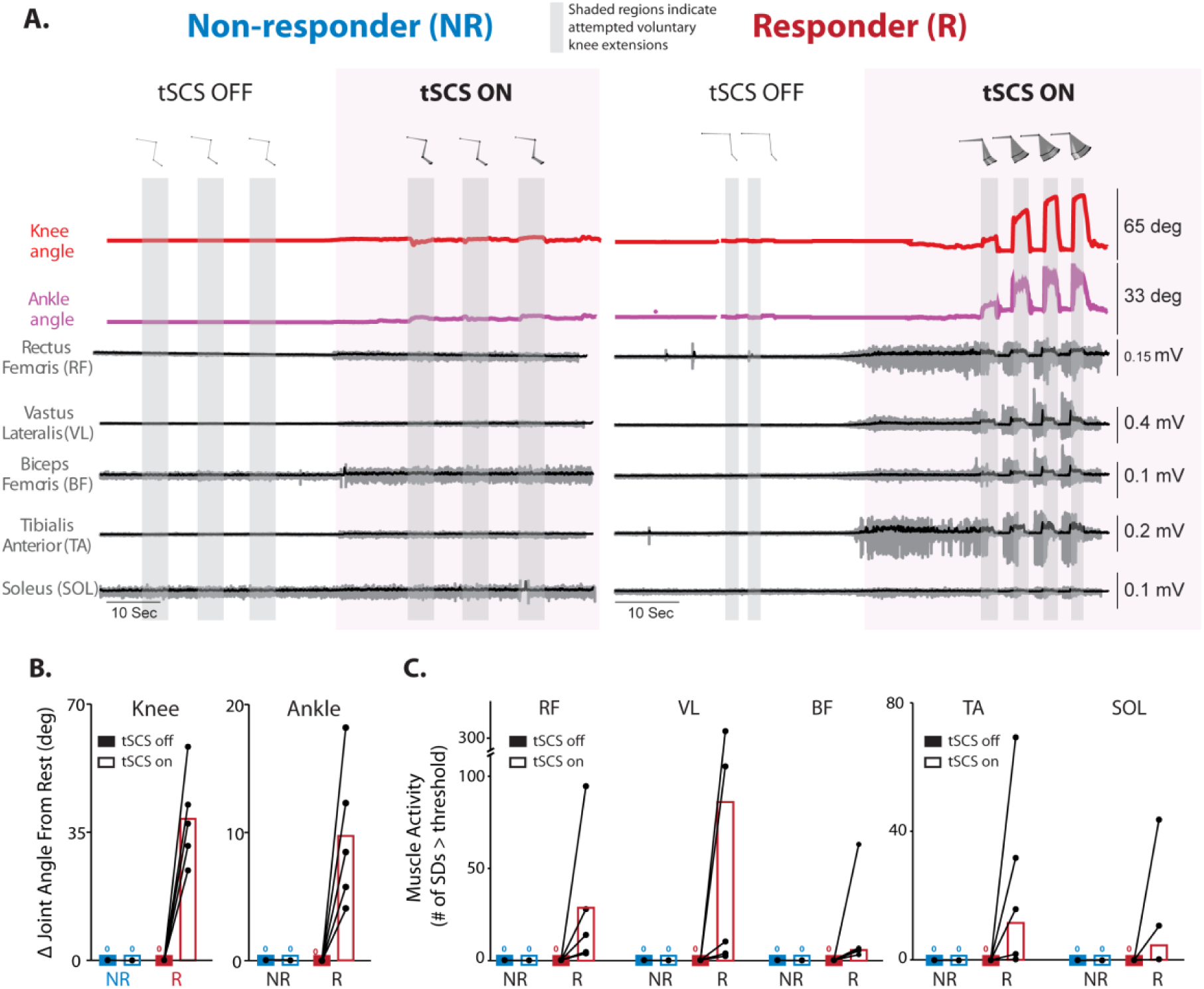
Kinematics and muscle activity of responders and non-responders. **A.** Joint angles and lower limb electromyography (EMG) of a non-responder (NR04 T5 AIS A) and responder (R01 (C4 AIS B). Grey shaded areas indicate periods of attempted contractions and purple shaded areas indicate when transcutaneous spinal cord stimulation (tSCS) was delivered. **B.** Change in knee and ankle angle from rest to attempted knee extension and dorsiflexion from a seated position in non-responders (blue) and responders (red) without tSCS (filled bars) and with tSCS (unfilled bars). The mean change in knee, and ankle angles during attempted rhythmic contract-relax cycles with tSCS in the responders was 38.6 ° ± 12.9, and 9.6 ° ± 5.6, respectively. **C.** Lower limb muscle activity in rectus femoris (RF), vastus lateralis (VL), biceps femoris (BF), tibialis anterior (TA), and soleus (SOL) without tSCS (filled bars) and with tSCS (unfilled bars) in non-responders (blue) and responders (red). EMG activity in RF, VL, BF, TA, and SOL exceeded the muscle activation threshold by a mean of 28.7 SDs ± 37.9, 85.9 SDs ± 132.1, 5.5 SDs ± 7.7, 11.5 SDs ± 29.6, 4.5 SDs ± 18.4, respectively. The muscle activation threshold was defined as the average background EMG activity (with or without tSCS) plus three standard deviations (see eMethods for further details). Muscle activity was quantified by the number of standard deviations exceeding the activation threshold.

*MEPs* induced from transcranial magnetic stimulation (TMS) of the primary motor cortex in the tibialis anterior (TA) were observed only in one of five responders (R03: latency: 52.5±0.7 ms, peak-to-peak amplitude at 100% stimulator output: 0.02±0.005mA) and no non-responders (Figure 2A). *Tibial nerve SEPs* were not observed in any participant (Figure 2B). *Soleus H-reflexes* were elicited in 4/5 responders and 2/5 non-responders (Figure 2C). In responders, the mean H_max_/M_max_ was 30±24% (Figure 2C), and in both NR02 and NR05 the ratio was 76% (group average, 30±41%). *Fibular CMAPs* were present in all participants except NR01 (Figure 2D). Considering the normative amplitude of a CMAP (3.0 mV)^20^, 4/5 responders showed normal responses (group average: 3.7±1.1mV), whereas only 2/5 non-responders showed normal amplitudes (group average, 2.4±1.7 mV) (Figure 2D). TA *muscle echo intensity* was outside the normative range (95% CI: 44.7 to 53.86) in all participants (Figure 2E). Individual data for each biomarker is in supplementary 1-5.

**Figure 2.**
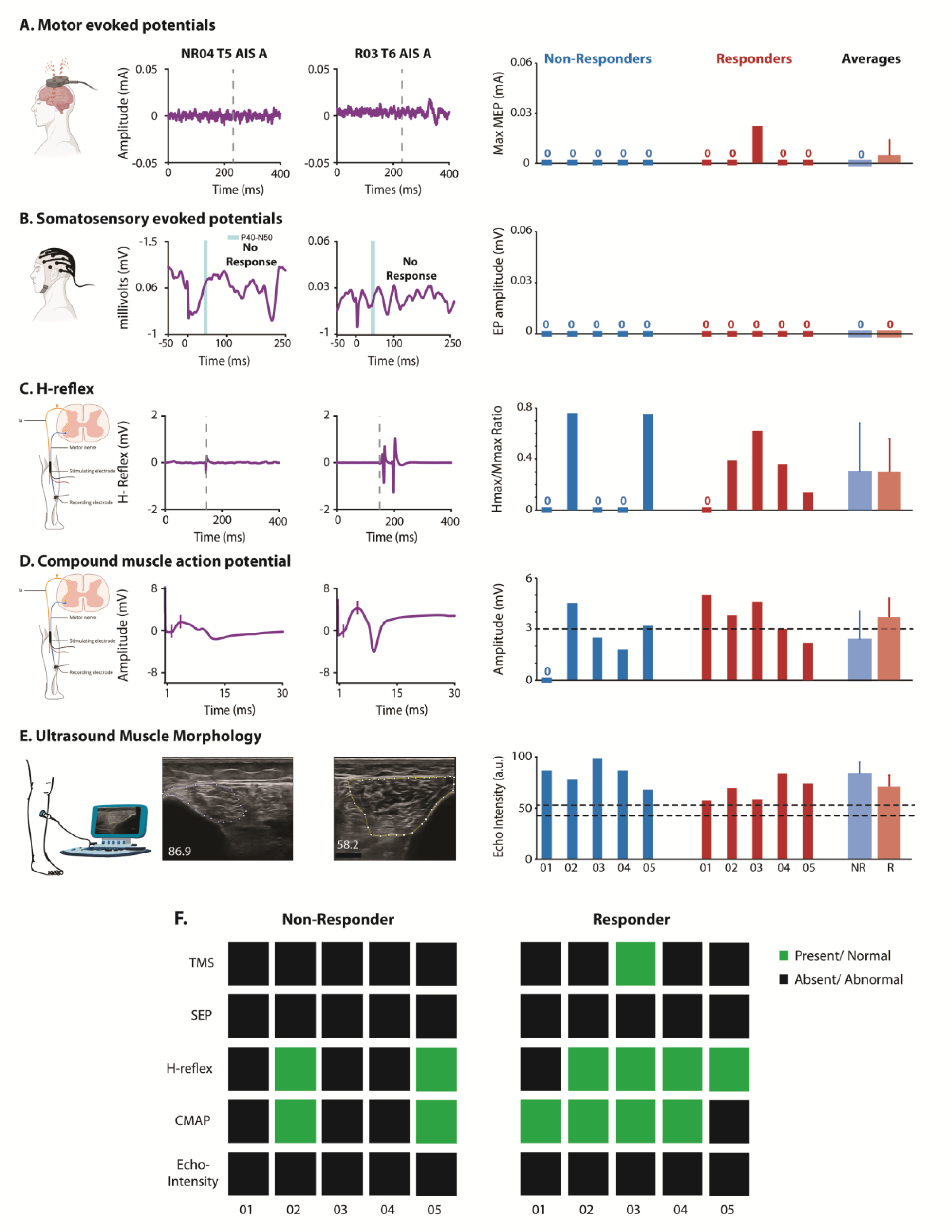
Neurophysiological and structural characteristics of responders and non-responders. For **A–E**, the left panels show individual data from a responder and non-responder, and the right panels show individual participant data and group averages. **A.** Recruitment curves for TMS-MEPs and quantified maximum MEPs without tSCS. **B.** Tibial nerve SEP traces and individual P40-N50 amplitudes. **C**. Soleus H-reflex and M-wave, and individual Hmax/Mmax ratio values. **D.** Fibular nerve CMAP waveforms and quantified amplitudes. Dotted lines represent normative CMAP amplitudes. **E.** Ultrasound images of the tibialis anterior and individual echo intensity values. Dotted lines indicate 95% confidence intervals from healthy controls. **F.** Summary table categorizing assessments as present/normal (green) or absent/abnormal (black) for non-responders and responders. **Abbreviations:** TMS; Transcranial Magnetic stimulation, SEP; Somatosensory Evoked Potential, H-Reflex; Hoffman Reflex, CMAP, Compound Muscle Action Potential.

## Discussion

We demonstrated that five individuals with cervical and upper thoracic motor-complete SCI (n = 3 AIS A, n = 2 AIS B) regained volitional knee extension and ankle dorsiflexion against gravity from a seated position during real-time tSCS after multi-week therapy. Previous research shows that tSCS facilitates oscillatory gait-like patterns in a gravity-eliminated position following motor-complete SCI (n = 5 AIS B) based on EMG and movement analyses.^2,4^ However, these studies did not report the return of volitional anti-gravity movements. While one study documented voluntary anti-gravity lower limb control using tSCS, it was limited to individuals with motor-complete SCI (n = 4, AIS B)^3^ and lacked quantitative data. This is the first study to use EMG and kinematics to demonstrate that tSCS can facilitate volitional lower limb movements against gravity in some individuals with AIS A and AIS B.

Responders were able to voluntarily extend their knee and dorsiflex their ankle in the presence of tSCS. This suggests that responders generated a supraspinal command to modulate the spinal circuitry controlling the legs. Based on this, we expected TMS over primary motor cortex to confirm enhanced corticospinal integrity in responders. However, only one responder (R03), and no other participant, showed TA MEPs (Figure 2). This aligns with prior studies that also reported absent TA and SOL MEPs induced by TMS in individuals with motor-complete SCI who regained voluntary movements with epidural SCS.^25,26^ Although MEPs have been detected in proximal and distal lower limb muscles following motor-complete SCI,^27^ they are rarely observed in injuries above T6. Our findings, combined with previous research, suggest that alternative mechanisms beyond cortico-motoneuronal connections—such as reticulospinal or cortico-reticulospinal pathways—may contribute to tSCS-mediated motor recovery.^28–30^

Of the assessments conducted, 4/5 responders exhibited H-reflexes and normal fibular CMAP amplitudes (Figure 2F), a trend suggesting that spinal cord reflex and peripheral motor nerve integrity are factors mediating responsiveness to tSCS. However, 2/5 non-responders also showed these responses, suggesting these metrics may lack specificity for distinguishing responders from non-responders to tSCS (Figure 2F). Despite this, the absence of an H-reflex and CMAP appears to be somewhat sensitive for detecting non-responsiveness (Figure 2F), highlighting their potential utility as screening biomarkers. Muscle morphology between responders and non-responders did not differ suggesting it may not be a good screening biomarker. While our data suggest that neurophysiological factors are likely important for responders in generating voluntary actions with tSCS, the residual descending pathway and important neuromuscular factors for restoring voluntary control of lower limb was not detected by the current measurements. Larger sample sizes and additional assessments of descending motor pathways beyond corticospinal are needed to identify specific markers to differentiate responders from non-responders.

Despite limitations such as sample size and variations in interventions, this study aimed to provide the first report of individuals with AIS A and AIS B who regained voluntary motor control using non-invasive tSCS at varying dosages, both with and without activity-based therapy. We developed a case series using responders from different clinical trials, as they were the only participants who regained voluntary movement across all trials. Non-responders were chosen to minimize the age-difference and the tSCS intervention used between groups. However, due to limited sample size we were unable to identify non-responders that underwent locomotor training alongside tSCS therapy. Our data shows that the return of voluntary movements can occur with and without activity-based rehabilitation. Future studies interventions with and without activity-based therapy are needed to determine the optimal strategy for recovery of voluntary movement control. In addition, future studies should consider additional neurophysiological and imaging assessments that were not included in this study. For instance, MRI could be utilized to assess white matter integrity, which has shown promise as a marker for motor responsiveness.^31,32^ TMS, combined with a startle stimulus, could also be used to probe reticulospinal excitability.^30,33^

In conclusion, tSCS enabled voluntary motor activity in five of 10 people with motor-complete SCI. Conventional neurophysiological measures were unable to identify pathways of motor recovery or differentiate responders from non-responders. Spinal cord reflex and peripheral motor nerve integrity may be important for responding to tSCS but may not distinguish responders from non-responders. Additional assessments and larger studies are needed to identify the pathways facilitating volitional control and the markers of motor responsiveness to tSCS.

## Acknowledgments

We would like to thank our study participants whose commitment to our research has made this project possible.

## Data availability

All data generated and analyzed during this study are included in this published article and in the online-only materials, further inquiries can be directed to the corresponding authors.

## Video Legends

** Please contact the corresponding author to request access to the videos

### Video 1: Responder 01 (R01)

This .mp4 video shows responder 01 (NLI; C3, AIS B) attempting simultaneous voluntary knee extension and ankle dorsiflexion in the absence (tSCS OFF) and presence (tSCS ON) of tSCS. **Abbreviations**: NLI, Neurological Level of Injury; AIS, American Spinal Injury Association Impairment Scale; tSCS, Transcutaneous Spinal Cord Stimulation.

### Video 2: Responder 02 (R02)

This .mp4 video shows responder 02 (NLI; T5, AIS A) attempting simultaneous voluntary knee extension and ankle dorsiflexion in the presence of tSCS (tSCS ON). **Abbreviations**: NLI, Neurological Level of Injury; AIS, American Spinal Injury Association Impairment Scale; tSCS, Transcutaneous Spinal Cord Stimulation.

### Video 3: Responder 03 (R03)

This .mp4 video shows responder 03 (NLI; T6, AIS A) attempting simultaneous voluntary knee extension and ankle dorsiflexion in the presence of tSCS (tSCS ON). **Abbreviations**: NLI, Neurological Level of Injury; AIS, American Spinal Injury Association Impairment Scale; tSCS, Transcutaneous Spinal Cord Stimulation.

### Video 4: Responder 04 (R04)

This .mp4 video shows responder 04 (NLI; T5, AIS A) attempting simultaneous voluntary knee extension and ankle dorsiflexion in the absence (tSCS OFF) and presence (tSCS ON) of tSCS. **Abbreviations**: NLI, Neurological Level of Injury; AIS, American Spinal Injury Association Impairment Scale; tSCS, Transcutaneous Spinal Cord Stimulation.

### Video 5: Responder 05 (R05)

This .mp4 video shows responder 05 (NLI; C5, AIS B) attempting simultaneous voluntary knee extension and ankle dorsiflexion in the absence (tSCS OFF) and presence (tSCS ON) of tSCS. **Abbreviations**: NLI, Neurological Level of Injury; AIS, American Spinal Injury Association Impairment Scale; tSCS, Transcutaneous Spinal Cord Stimulation.

