## Supplemental Figures for "Non-invasive spinal cord neuromodulation enables volitional anti-gravity leg movements after motor-complete spinal cord injury: responders vs. non-responders"

### Supplemental Online-Only Figures

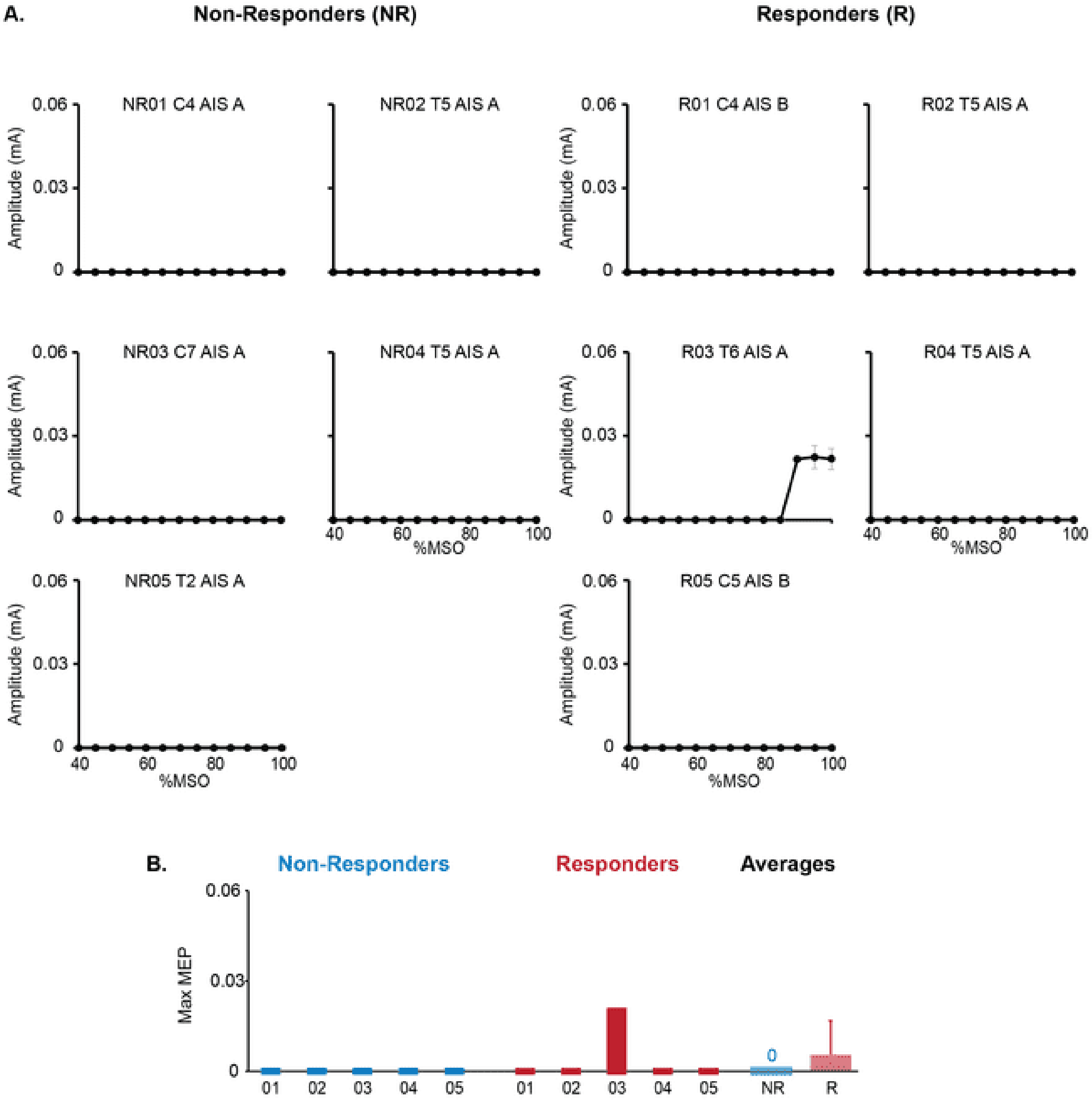


**Supplemental Figure 1: Motor evoked potentials**
**(A)** Recruitment curves of motor-evoked potentials in the Tibialis Anterior, elicited by Transcranial Magnetic Stimulation over the motor cortex for each responder and non-responder. **(B)** The maximum MEP amplitude for each participant and group averages with tSCS off.

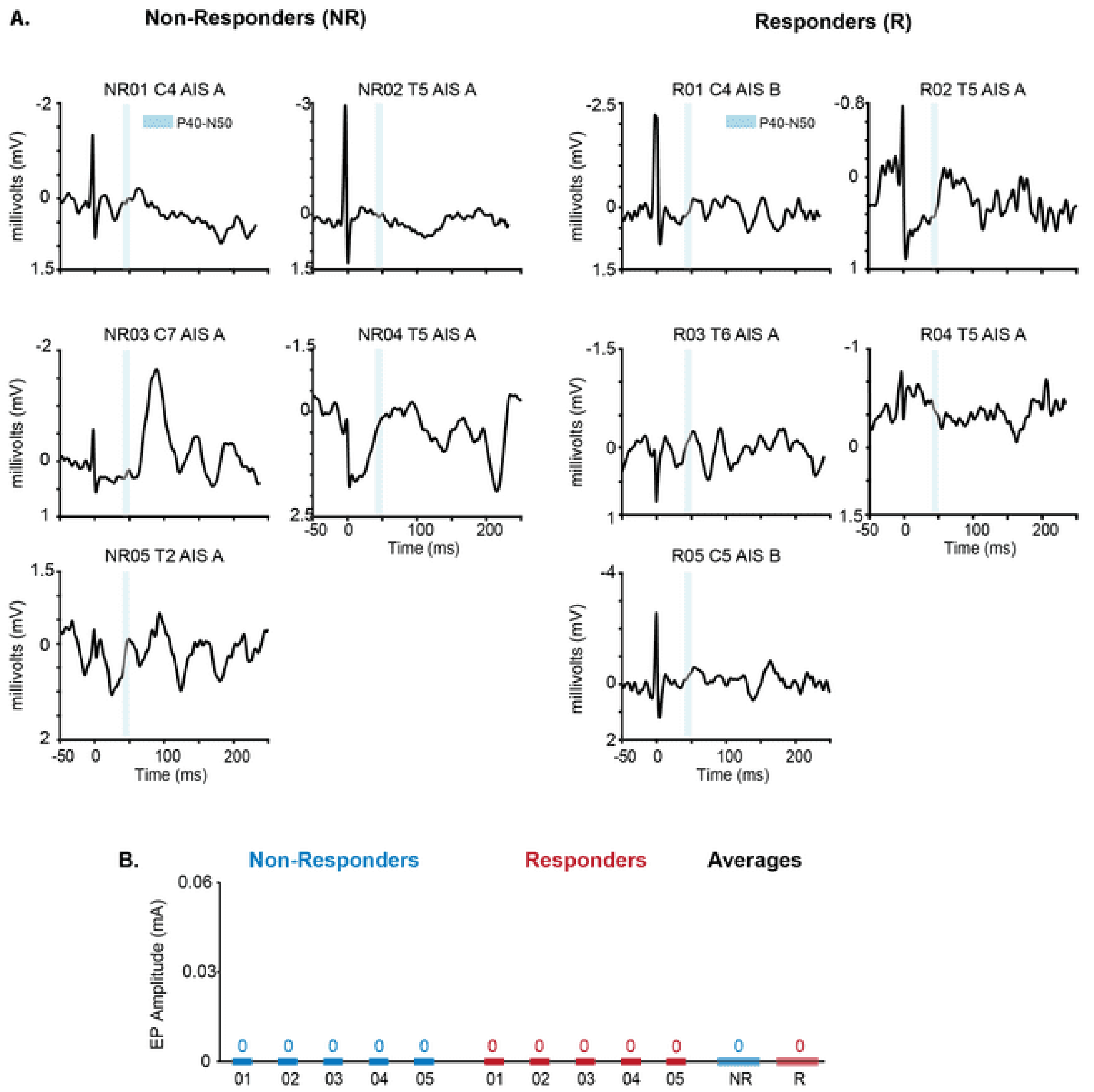


**Supplemental Figure 2: Somatosensory evoked potentials**

**(A)** Tibial nerve SEP traces and P40-N50 amplitudes (blue shaded region) recorded from Cz referenced to Fz for each responder (R) and non-responder (NR). **(B)** P40-N50 amplitude for each participant and group averages with no spinal cord stimulation.

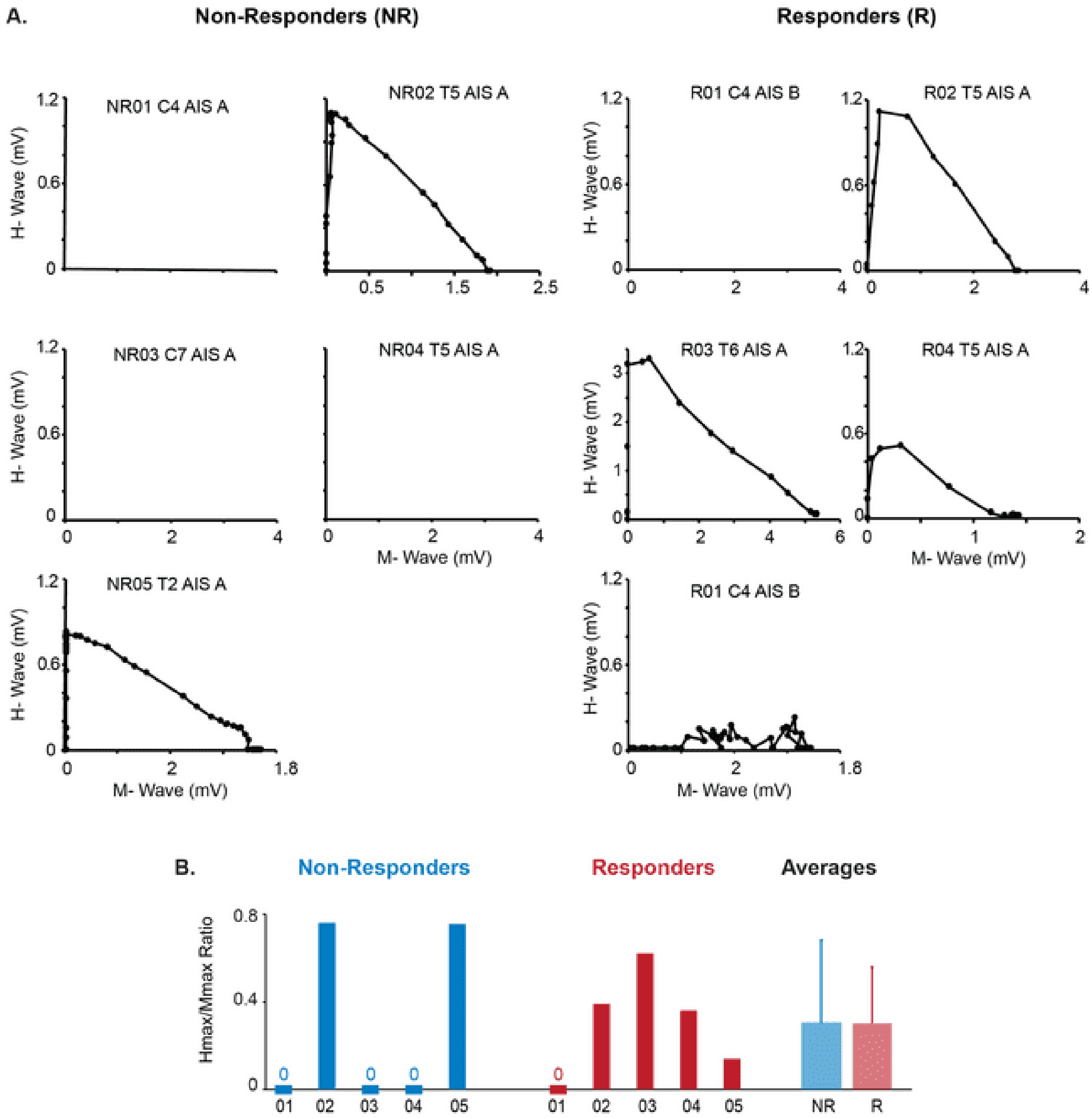


**Supplemental Figure 3: Hoffman (H) reflex**

**(A)** Soleus H-reflex versus M-wave for each responder (R) and non-responder (NR). **(B)** Hmax/Mmax ratio for each participant and group averages with with no spinal cord stimulation.

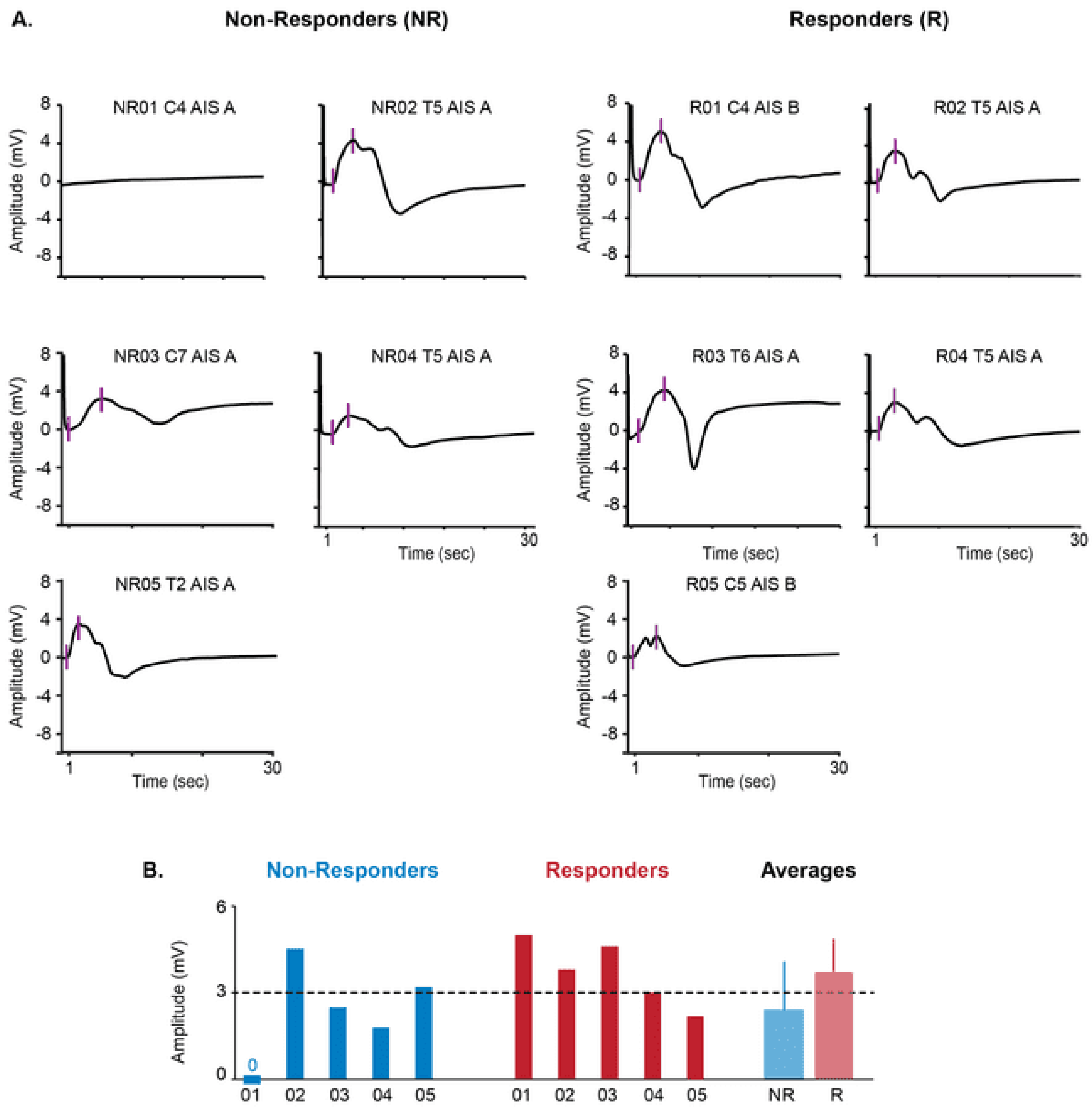


**Supplemental Figure 4: Compound muscle action potentials (CMAPs)**
**(A)** Fibular nerve CMAP waveforms for each responder and non-responder. **(B)** Quantified CMAP amplitudes for each participant and group averages with no spinal cord stimulation. Dotted line represents normative amplitudes.

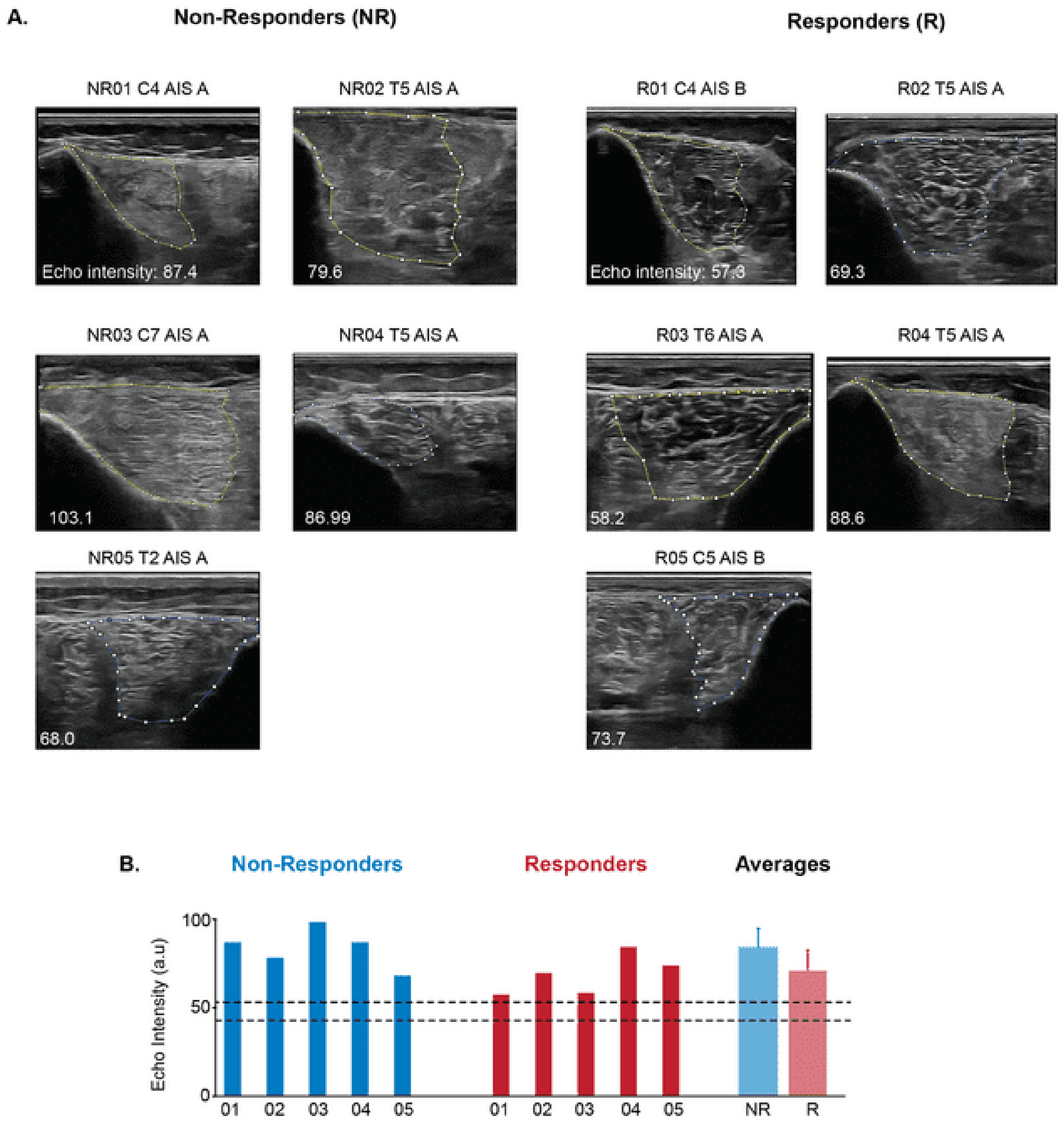


**Supplemental Figure 5: Muscle morphology**

**(A)** Ultrasound images of the tibialis anterior for each responder and non-responder. **(B)** Quantified echo-intensity values for each participant and group averages with tSCS off. Dotted lines indicate 95% confidence intervals from healthy controls (see Methods).
